# Neutralization of ancestral SARS-CoV-2 and variants Alpha, Beta, Gamma, Delta, Zeta and Omicron by mRNA vaccination and infection-derived immunity through homologous and heterologous variants

**DOI:** 10.1101/2021.12.28.21268491

**Authors:** Meriem Bekliz, Kenneth Adea, Pauline Vetter, Christiane S Eberhardt, Krisztina Hosszu-Fellous, Diem-Lan Vu, Olha Puhach, Manel Essaidi-Laziosi, Sophie Waldvogel-Abramowski, Caroline Stephan, Arnaud G. L’Huillier, Claire-Anne Siegrist, Arnaud M Didierlaurent, Laurent Kaiser, Benjamin Meyer, Isabella Eckerle

## Abstract

Emerging SARS-CoV-2 variants of concern/interest (VOC/VOI) raise questions about effectiveness of neutralizing antibodies derived from infection or vaccination. As the population immunity to SARS-CoV-2 has become more complex due to prior infection and/or vaccination, understanding the antigenic relationship between variants is needed.

Here, we have assessed in total 104 blood specimens from convalescent individuals after infection with early-pandemic SARS-CoV-2 (pre-VOC) or with Alpha, Beta, Gamma or Delta, post-vaccination after double-dose mRNA-vaccination and break through infections due to Delta or Omicron. Neutralization against seven authentic SARS-CoV-2 isolates (B.1, Alpha, Beta, Gamma, Delta, Zeta, Omicron) was assessed by plaque-reduction neutralization assay.

We found highest neutralization titers against the homologous (previously infecting) variant, with lower neutralization efficiency against heterologous variants. Significant loss of neutralization for Omicron was observed but to a varying degree depending on previously infecting variant (23.0-fold in Beta-convalescence up to 56.1-fold in Alpha-convalescence), suggesting that infection-derived immunity varies, but independent of the infecting variant is only poorly protective against Omicron. Of note, Zeta VOI showed also pronounced escape from neutralization of up to 28.2-fold in Alpha convalescent samples. Antigenic mapping reveals both Zeta and Omicron as separate antigenic clusters.

Double dose vaccination showed robust neutralization for Alpha, Beta, Gamma, Delta and Zeta, with fold-change reduction of only 2.8 (for Alpha) up to 6.9 (for Beta). Escape from neutralization for Zeta was largely restored in vaccinated individuals, while Omicron still showed a loss of neutralization of 85.7-fold compared to pre-VOC SARS-CoV-2.

Combined immunity from infection followed by vaccination or vaccine breakthrough infection showed highest titers and most robust neutralization for heterologous variants. Breakthrough infection with Delta showed only 12.5-fold reduced neutralization for Omicron, while breakthrough infection with Omicron showed only a 1.5-fold loss for Delta, suggests that infection with antigenically different variants can boost immunity for antigens closer to the vaccine strain. Antigenic cartography showed also a tendency towards broader neutralizing capacity for heterologous variants.

We conclude that the complexity of background immunity needs to be taken into account when assessing new VOCs. Development towards separate serotypes such as Zeta was already observed before Omicron emergence, thus other factors than just immune escape must contribute to Omicrons rapid dominance. However, combined infection/vaccination immunity could ultimately lead to broad neutralizing capacity also against non-homologous variants.

## Introduction

In late 2019, the Severe Acute Respiratory Syndrome Coronavirus 2 (SARS-CoV-2) emerged in Wuhan (China) causing a pandemic that led to an unprecedented international health crisis of yet unknown outcome [1, 2]. Shortly after its emergence, SARS-CoV-2 acquired the D614G mutation in the spike protein in February 2020, which quickly replaced all other circulating variants and spread worldwide. The evolutionary advantage of D614G is associated with enhanced binding to the human receptor and increased replication and thus presumably better transmissibility [3]. After largely uncontrolled transmission on a global scale during 2020, the emergence of the first variants of concern (VOCs) was observed [4]. The VOCs consist of the Alpha variant, first detected in the UK; the Beta variant, first detected in South Africa, the Gamma variant, first detected in South America, the Delta variant, first detected in India and the Omicron variant, very recently reported from South Africa [5]. Variants of concern were characterized by a rapid increase in case numbers and they quickly outcompeted earlier strains in their region of emergence. The latest emergence is the Omicron variant in late 2021 with the so far highest number of mutations compared to earlier variants and the majority of them located in the spike protein [4]. Thus, suspicion of escape from antibody responses derived from earlier variants and vaccines by Omicron is high.

In addition to VOCs, other variants were identified that were of less concern and therefore classified as variants of interest (VOI) due to aspects in their epidemiology or genetic signatures potentially leading to an altered phenotype, among them the (former) VOI Zeta that arose in parallel with the Gamma variant in South America at a time when a local surge in cases was observed but has disappeared in the meantime [6-10].

Currently, few treatments are widely available for SARS-CoV-2 and mostly dedicated to risk groups, therefore prevention and protection through vaccine-mediated immunity is still the proposed method for ending the pandemic [11]. Depending on countries, medium to high levels of population immunity have already been reached through vaccination or infection, but there are huge geographical differences when it comes to the proportions of the population infected, different circulation of variants against which immunity was obtained, percentage of vaccinated individuals, vaccine doses and type of vaccine used. In light of that, it is of particular importance to evaluate the neutralizing potential of elicited antibodies against clinical isolates of VOCs/VOIs to detect immune escape variants early and understand the impact of such variants on the further course of the pandemic.

The mRNA-based vaccine Pfizer-BioNTech BNT162b2 encodes a stabilized full-length SARS-CoV-2 spike ectodomain derived from the Wuhan-Hu-1 genetic sequence and elicits potent neutralizing antibodies [12], as does the mRNA-based vaccine Moderna mRNA-1273 [13]. However, emerging SARS-CoV-2 variants include multiple substitutions and deletions in the major target of neutralizing antibodies, the spike glycoprotein, including the N-terminal (NTD) and the receptor-binding domains (RBD), with the largest number of mutations of over 30 observed in the Omicron variant. This raises the question of whether neutralizing antibodies induced by early circulating strains or by current vaccines can effectively neutralize recently emerged virus variants. So far, studies indicated that mutations that have been accumulating in the spike protein, especially in the RBD, are associated with increased affinity to the human ACE2 receptor [14], as well as with resistance to neutralization from antibodies of previously infected or vaccinated patients [15]. Mutations in the RBD pose the greatest risk for making SARS-CoV-2 more infectious and able to escape antibody neutralization [14]. Virus neutralizing antibody have been shown to be a correlate of protection from SARS-CoV-2 but more insights on neutralizing responses against emerging virus variants are needed [16-19].

In this study, we investigated the neutralizing potency of a panel of authentic sera or plasma from individuals vaccinated twice with either BNT162b2 or mRNA-1273, and convalescent patients that had mild coronavirus disease 2019 (COVID-19). Patients were infected at different time points during the pandemic with either an early-pandemic (pre-VOC) SARS-CoV-2 or one of the VOCs Alpha, Beta, Gamma, Delta or Omicron. We used authentic clinical isolates for pre-VOC SARS-CoV-2 (Pangolin lineage B.1), Alpha, Beta, Gamma, Delta, Zeta and Omicron which were all isolated from patient samples collected from our routine diagnostic laboratory. We assessed the neutralizing potential against homologous and heterologous variants by live virus plaque reduction neutralization test (PRNT), widely regarded as the gold standard for the detection of SARS-CoV-2-specific neutralizing antibodies [20].

## Materials and Methods

### Setting

The laboratory of virology of the University Hospital of Geneva is participating and coordinating the SARS-CoV-2 variant and genomic surveillance funded by the Swiss Federal Office of Public Health with constantly ongoing full genome sequencing of SARS-CoV-2 positive patient samples obtained through the diagnostic unit of our Centre [21]. Up to around 400 positive patient specimens with a cycle threshold (Ct) < 32 are sequenced each week since march 2021. From each variant that falls into any of the categories of variant of concern/interest, at least one virus isolate is generated.

### Patient samples

Convalescent sera or plasma during the early pandemic period (pre-VOC) were collected in the context of two prospective cohort studies at the Geneva University Hospitals (HUG) and the Geneva Centre for Emerging Viral Diseases (Understanding COVID study; ethics approval number: CCER 2020-00516, Persistence study: ethics approval number: CCER 2020-00516). In addition, anonymized left-over samples from apheresis collection of plasma (all collected in 2020) were available under the general informed consent of the University Hospitals of Geneva.

Convalescent sera obtained from patients infected with a SARS-CoV-2 variant of concern (Alpha, Beta, Gamma, Delta, Omicron) were collected in 2021 by contacting patients with confirmed SARS-CoV-2 infection for a blood collection in the convalescent period for the purpose of this study, same for vaccine breakthrough infections (Ethics approval number: CCER 2020-02323). For each patient infected with a variant, information on the infecting virus was available by full-genome sequence. No sequence information on the infecting strain was available from patients infected in 2020, however the first SARS-CoV-2 variant in Switzerland (Alpha) was only observed on Dec 24, 2020, and all pre-VOC samples were collected before that date, thus they are considered pre-VOC. Plasma samples from vaccinated healthy individuals, vaccinated with 2 doses of BNT162b2 (Pfizer/BioNTech) or mRNA-1273 (Moderna) vaccine at 28 days interval were available from a prospective observational study, collected 30 days after the 2^nd^ dose (Ethics approval number: CCER ICOVax 2021-00430).

Convalescent samples were only collected from individuals with RT-PCR-confirmed diagnosis of SARS-CoV-2 in our diagnostic unit, and sera/plasma were collected 3-137 days after diagnosis or symptom onset (days post diagnosis, DPP). All vaccinated individuals were in addition tested for the presence of nucleocapsid antibodies (Elecsys® Anti-SARS-CoV-2 anti-N) to screen for unrecognized infection prior to vaccination. There are no differences to be expected in the PRNT regarding the use of plasma or serum, therefore both sample types are used in parallel and termed “convalescent samples” throughout the manuscript.

### Viruses and cells

Vero-E6 and Vero E6-TMPRSS cells were cultured in complete DMEM GlutaMax I medium supplemented with 10% fetal bovine serum, 1x Non-essential Amino Acids, and 1% antibiotics (Penicillin/Streptomycin) (all reagents from Gibco, USA). Vero-TMPRSS were kindly received from National Institute for Biological Standards and Controls (NIBSC, Cat. Nr. 100978).

All SARS-CoV-2 viruses used in this study were isolated from residual nasopharyngeal swabs collected from patients presenting at the Geneva University Hospitals under general informed consent of the hospital that allows usage of anonymized left-over materials. All patient specimens from which isolates were obtained were fully sequenced (**Table S1**). The following viruses were isolated as follows: B.1, Alpha, Gamma and Zeta were isolated and propagated on Vero E6. The Beta variant isolate was obtained as described previously [22]. Briefly, no primary Beta isolate could be obtained on VeroE6 but only on A549 cells overexpressing human ACE2 [23]. Therefore, after primary isolation, A549-hACE2 cells were mixed with VeroE6 in a 1:1 ratio and inoculated with the passage 1 isolate. The next passage was done on VeroE6 to generate the virus stock. The Omicron variant was primarily isolated on Vero-TMPRSS cells, then transferred to Vero E6 for generation of a virus stock. All virus stocks were titrated on Vero-E6 cells and full genome sequenced. Sequences of initial patient specimens and virus isolates obtained were compared for acquired mutations. Experiments with live infectious SARS-CoV-2 followed the approved standard operating procedures of our biosafety level 3 facility (BSL-3).

### Plaque Reduction Neutralization Test (PRNT)

Following the PRNT procedure, Vero-E6 cells were seeded at a density of 4×10^5^ cells/mL in 24-well cell culture plates. A total of 34, 12, 8, 10 and 22 sera/plasma samples from patients infected with pre-VOC SARS-CoV-2, Alpha, Beta Gamma, Delta or sera from individuals vaccinated with BNT162b2/mRNA-1273, respectively, were used for determining the neutralizing titers against B.1 (first pandemic wave strain containing only the D614G substitution in the spike gene), Alpha, Beta, Gamma, Zeta, Delta and Omicron variants as described earlier [24]. Briefly, all sera/plasma were heat-inactivated at 56°C for 30 min and serially diluted in Opti-Pro serum-free medium starting from 1:10 until up to 1:5120 if necessary. Sera/plasma were mixed with 50PFU of variant isolates (B.1, Alpha, Beta, Gamma, Delta, Zeta and Omicron) and incubated at 37°C for 1h. All samples were run in duplicate and for each neutralization experiment an infection control (no serum/plasma) and a reference serum were used to ensure reproducibility between different experiments. Vero-E6 cells were washed 1x with PBS and inoculated with the virus serum/plasma mixture for 1h. Afterwards, the inoculum was removed and 500uL of the overlay medium also used for the plaque assays was added. After incubation for 3 days at 37°C, 5% CO2, the overlay medium was removed, cells were fixed in 6% formaldehyde solution for at least 1h, plates were washed 1x with PBS and stained with crystal violet. Plaques were counted in wells inoculated with virus-serum/plasma mixtures and compared to plaque counts in infection control wells. The 90% reduction endpoint titers (PRNT_90_) were calculated by fitting a 4-parameter logistics curve with variable slope to the plaque counts of each serum/plasma using GraphPad Prism version 9.2.0. For samples that did not reach 90% reduction at a 1:10 dilution, we extrapolated the titer until a dilution of 1, i.e. undiluted sample. If the extrapolation reached a titer below 1, the sample was given a value of 0.5 and considered negative.

### Antigenic cartography

Antigenic maps were constructed with the software from https://acmacs-web.antigenic-cartography.org. The maps were generated with PRNT90 titers obtained for convalescence specimens from pre-VOC SARS-CoV-2, Alpha, Beta, Gamma and Delta and for post-vaccine specimens with and without prior infection, as described previously [25]. Breakthrough infections with Delta and Omicron were not included due to low number of samples.

### Statistical analysis

Geometric means with 95% CI were used for comparison of PRNT_90_ titers. All statistical analyses were conducted using GraphPad Prism version 9.1.0 software, performed using log10 ANOVA transformed PRNT_90_ titers. Results were considered significant with p values <0.05.

## Results

To evaluate the neutralization capacity of SARS-CoV-2 specific antibodies against SARS-CoV-2 variants (pre-VOC, Alpha, Beta, Gamma, Delta, Zeta and Omicron), a panel of convalescent blood specimens were used from i) individuals previously infected with pre-VOC SARS-CoV-2 (n=34), ii) individuals previously infected with VOCs Alpha (n=12), Beta (n=8), Gamma (n=10), Delta (n=10) iii) BNT162b2 or mRNA-1273 vaccinated individuals with (n=6) and without prior infection (n=16) iv) BNT162b2 or mRNA-1273 vaccinated individuals with a break-through infection with either Delta (n=4) or Omicron (n=4) (**Table 1**).

**Table 1.** Characteristic of sample cohorts used in this study. DPP, days post diagnosis, DPOS, days post symptom onset (marked with *) WPV, weeks post 2nd dose vaccination, n.a. not accessible

| **Convalescent cohort** |  |  |  |  |  |  |
| --- | --- | --- | --- | --- | --- | --- |
| **Infecting virus** | **Number patient** | **Gender (M/F)** | **Age, mean** | **DPP/DPOS** | **Sample type** | **Infection period** |
| **pre-VOCs** | 34 | 20/14 | 31 | 32 (25-37)* | Serum, Plasma | March-June 2020 |
| **Alpha** | 12 | 8/4 | 51 | 27 (8-42) | Serum | December 2020-February 2021 |
| **Beta** | 8 | 2/6 | 42 | 50 (3-98) | Serum | January-May 2021 |
| **Gamma** | 10 | 4/6 | 44 | 54 (7-137) | Serum | January-April 2021 |
| **Delta** | 10 | 4/6 | 42 | 71 (9-118) | Serum | May-July 2021 |
| **Vaccinated cohort** |  |  |  |  |  |  |
| **Vaccine** | **Number of patient** | **Gender (M/F)** | **Age mean** | **WPV Mean weeks (range)** | **Sample type** | **Date of vaccination** |
| **mRNA vaccine** | 16 | 4/12 | 52 | 8 | Serum, Plasma | March-May 2021 |
| **Vaccinated and infected individuals** |  |  |  |  |  |  |
| **Vaccination/infection status** | **Number of patient** | **Gender (M/F)** | **Age mean** | **WPV Mean weeks (range)** | **Sample type** | **Intervall vaccination – infection**<br>**Mean weeks (range)** |
| **Prior infection + 2x vaccination** | 6 | 3/3 | 46 | n.a | Serum | n.a. |
| **2x vaccination + breakthrough (Delta)** | 4 | 2/2 | 42 | 28 (16-37) | Serum | 20 (11-28) |
| **2x vaccination + breakthrough (Omicron)** | 2 | 1/1 | 49 | 26 (24-28) | Serum | 23 (21-25) |
| **1x vaccination + breakthrough (Omicron)** | 2 | 2/0 | 13 | 5 (5-5) | Serum | 3 (3) |

### Neutralizing capacity from infection-derived convalescent samples

Geometric mean PRNT_90_ titers of convalescent specimens from individuals infected with pre-VOC SARS-CoV-2 were 37.3 (95%CI: 25.4-54.9) against B.1, 16.7 (95%CI: 9.7-28.8) against Alpha, 14.0 (95%CI: 8.8-22.3) against Beta, 10.3 (95%CI:6.4-16.6) against Gamma, 12.0 (95%CI:7.0-20.3) against Delta, 1.4 (95%CI: 0.9-2.4) against Zeta, and 0.8 (95%CI:0.6-1.2) against Omicron. Compared to B.1 this results in a fold-change reduction of 2.2 for Alpha, 2.7 for Beta, 3.6 for Gamma, 3.1 for Delta, 25.9 for Zeta and 45.6 for Omicron (**Figure 1A**). None of the samples failed to neutralize the homologous virus. Only 1/34 (3%), 2/34 (6%), 3/34 (9%) and 3/34 (9%) failed to neutralize Alpha, Beta, Gamma and Delta, respectively, whereas 21/34 (62%) and 29/34 (85%) completely failed to neutralize the Zeta and Omicron variant.

**Figure 1.**
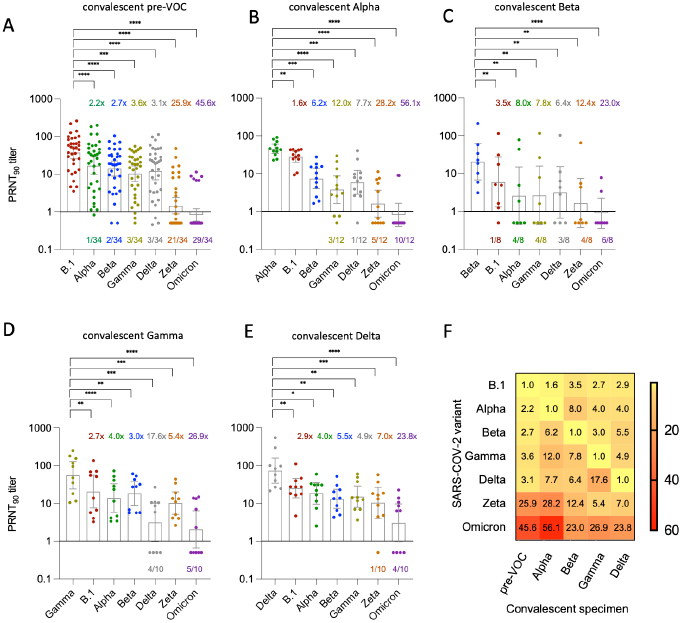
Neutralization in infection-derived blood specimens against seven authentic isolates of SARS-CoV-2 variants (B.1, Alpha, Beta Gamma, Delta, Zeta, Omicron). Bars represent geometric mean titers (GMT) of 90% reduction endpoint titers (PRNT_90_) with 95% confidence interval. Convalescent specimens are derived from individuals infected with (**A**) early-pandemic SARS-CoV-2 (pre-VOC), (**B**) Alpha (**C)** Beta (**D**) Gamma (**E**) Delta. Colored numbers above bars refer to fold change reduction of GMT versus the homologous (infecting) variant, shown as first bar of each figure. Colored numbers below each bar represents number of specimens with complete loss of neutralization (PRNT_90_ titer < 1). *p < 0.05, **p <0.003, ***p<0.0002 and ****p < 0.0001 (**F**) Heatmap of fold-reduction in neutralization based on PRNT_90_ data from A-E.

For patients previously infected with the Alpha variant (n=12), geometric mean PRNT_90_ titers were 45.5 (95%CI:34.3-60) for Alpha, 27.8 (95%CI:19.8-39.0) for B.1, 7.4 (95%CI:4.0-13.5) for Beta, 3.8 (95%CI:1.6-8.7) for Gamma, 5.9 (95%CI:2.8-12.2) for Delta, 1.6 (95%CI:0.7-3.6) for Zeta and 0.8 (95%CI:0.4-1.7) for Omicron. Compared to the homologous Alpha variant, this results in a reduction of 1.6 (B.1), 6.2 (Beta), 12.0 (Gamma), 7.7 (Delta), 28.2 (Zeta) and 56.1 (Omicron). Complete loss of neutralization was observed for 3/12 (25%) samples for Gamma, 1/12 (8%) for Delta, 5/12 (42%) for Zeta and 10/12 (83%) for Omicron (**Fig. 1B**).

For individuals previously infected with the Beta variant (n=8), geometric mean PRNT_90_ titers were 20.6 (95%CI:6.8-62.6) for Beta, 6.0 (95%CI:1.3-27.2) for B.1, 2.6 (95%CI:0.4-14.9) for Alpha, 2.6 (95%CI:0.4-15.8) for Gamma, 3.2 (95%CI:0.7-15.3) for Delta, 1.7 (95%CI:0.4-7.5) for Zeta and 0.9 (95%CI:0.4-2.3) for Omicron. Compared to the homologous virus (Beta), this results in a fold-reduction of 3.5 for B.1, 8.0 for Alpha, 7.8 for Gamma, 6.4 for Delta, 12.4 for Zeta and 23.0 for Omicron. Complete loss of neutralization was observed for 1/8 (12.5) for B.1, 4/8 (50%) for Alpha, 4/8 (50%) for Gamma, 3/8 (37.5) for Delta, 4/8 (50%) for Zeta and 6/8 (75%) for Omicron (**Fig. 1C**).

For individuals previously infected with the Gamma variant (n=10), geometric mean PRNT_90_ titers were 55.6 (95%CI:24.1-128) for Gamma, 20.5 (95%CI:7.6-55.5) for B.1, 13.9 (95%CI:5.9-32.9) for Alpha, 18.3 (95%CI:8.9-37.4) for Beta, 3.2 (95%CI:1.0-10.1) for Delta, 10.2 (95%CI:5.2-20.2) for Zeta and 2.1 (95%CI:0.7-6.4) for Omicron. Compared to the homologous virus, fold reduction in neutralization was 2.7 for B.1, 4.0 for Alpha, 3.0 for Beta, 17.6 for Delta, 5.4 for Zeta and 26.9 for Omicron. Complete loss of neutralization was observed for 4/10 (40%) for Delta and 5/10 (50%) for Omicron (**Fig. 1D**). Of note, a rather strong loss of neutralization in Gamma convalescent samples was observed for Delta, while neutralization was less affected for the Zeta variant.

For individuals previously infected with Delta (n=10), geometric mean PRNT_90_ titers were 72.8 (95%CI:33.9-156.2) for Delta, 25.1 (95%CI:14.0-45.1) for B.1, 18.4 (95%CI:9.5-35.4) for Alpha, 13.2 (95%CI:7.4-23.5) for Beta, 15.0 (95%CI:7.8-28.7) for Gamma, 10.4 (95%CI:4.0-26.9) for Zeta and 3.1 (95%CI:1.0-9.6) for Omicron. Fold reduction compared to homologous virus (Delta) was 2.9 for B.1, 4.0 for Alpha, 5.5 for Beta, 4.9 for Gamma, 7.0 for Zeta and 23.8 for Omicron. Complete loss of neutralization was 1/10 (10%) for Zeta and 4/10 (40%) for Omicron (**Fig. 1E**).

A heatmap for fold-change reduction of neutralization was generated to summarize findings across all convalescent specimens (**Fig. 1F**). Here, rather robust neutralization of pre-VOC convalescent specimens against VOCs Alpha, Beta, Gamma and Delta is visible while the other variants showed stronger escape from immunity against heterologous variant. Immune escape properties for Zeta and Omicron are visible, with stronger immune escape of Zeta from pre-VOC and Alpha immunity, but less from Gamma and Delta immunity. Immune escape of Omicron was pronounced throughout all specimens, although the fold-change reduction is in a comparable range to that of Zeta for some combinations.

### Neutralizing capacity from post-vaccine and combined post-vaccine/infection-derived samples

We investigated a total of 30 patient specimens from either double-vaccinated individuals (n=16) or with combined vaccination-infection-derived immunity, either through prior infection followed by vaccination, or vaccination followed by a vaccine breakthrough infection with Delta or Omicron (n=14).

In contrast to all infection-derived convalescent samples, geometric mean PRNT_90_ titers were much higher for individuals double-vaccinated with either BNT162b2 or mRNA-1273 with titers of 338.0 (95%CI:247.4-461.6) against B.1, 121.7 (95%CI:86.0-172.3) against Alpha, 49.3 (95%CI:28.1-86.8) for Beta, 62.8 (95%CI:36.0-109.5) for Gamma, 95.6 (95%CI:69.4-131.7) for Delta, 78.5 (95%CI:50.4-122.5) for Zeta and 3.9 (95%CI:1.8-8.7) for Omicron. This translates into a fold reduction of neutralization of 2.8 for Alpha, 6.9 for Beta, 5.4 for Gamma, 3.5 for Delta, 4.3 for Zeta and 85.7 for Omicron. No complete loss of neutralization was seen for any VOC except Omicron in 5/16 (31%) specimens (**Fig. 2A**).

**Figure 2.**
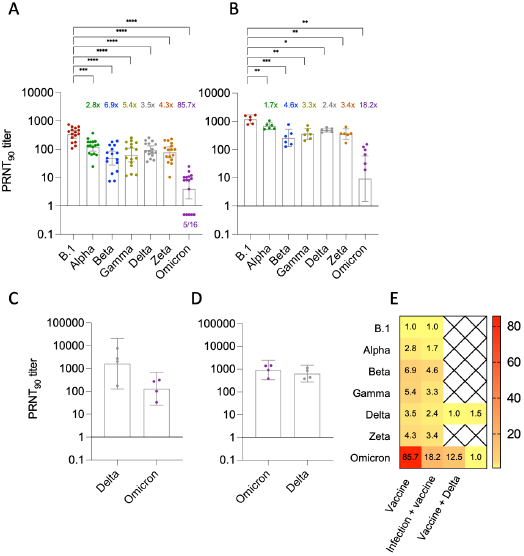
Neutralization in infection-derived blood specimens against seven authentic isolates of SARS-CoV-2 variants (B.1, Alpha, Beta Gamma, Delta, Zeta, Omicron). Bars represent geometric mean titers (GMT) of 90% reduction endpoint titers (PRNT_90_) with 95% confidence interval. Cohorts consist of individuals with (**A**) double-dose mRNA vaccination, (**B**) prior SARS-CoV-2 infection followed by double-dose mRNA vaccination (**C**) Delta breakthrough infection of double-vaccinated individuals and (**D**) Omicron breakthrough infection following double (n=2) and single (n=2) mRNA vaccination. Colored numbers above bars refer to fold change reduction of GMT versus the homologous (infecting) variant, shown as first bar of each figure. Colored numbers below each bar represents number of specimens with complete loss of neutralization (PRNT_90_ titer < 1). *p < 0.05, **p <0.003, ***p<0.0002 and ****p < 0.0001 (**F**) Heatmap of fold-reduction in neutralization based on PRNT_90_ data from A-D.

Individuals with prior SARS-CoV-2 infection before double vaccination (n=6), as determined by presence of antibodies against the nucleocapsid, showed geometric mean PRNT_90_ titers of 1190.4 (95%CI:837.8-1691) against B.1, followed by 683.2 (95%CI:516.3-904.1) for Alpha, 260.4 (95%CI:128.8-526.5) for Beta, 360.4 (95%CI:224.5-578.5) for Gamma, 494.1 (95%CI:419.4-582.1) for Delta, 351.8 (95%CI:227-545.2) for Zeta and 65.2 (95%CI:27.81-153.0) for Omicron. Fold reduction in neutralization was 1.7 for Alpha, 4.6 for Beta, 3.3 for Gamma, 2.4 for Delta, 3.4 for Zeta and 18.2 for Omicron. Of note, none of the specimens showed complete loss of neutralization (**Fig. 2B**).

In addition, we have investigated vaccinated individuals with a breakthrough infection with Delta (n=4) and Omicron (n=4) for neutralization against both viruses which are currently the only VOCs co-circulating. For the first group, high geometric mean PRNT_90_ titers of 1636 (95%CI: 128.2-20885) were observed for the Delta while 130.9 (95%CI:24.9-688.9) was observed for Omicron. This results in a 12.5-fold reduction versus the homologous Delta, but no complete loss of neutralization was observed (**Fig. 2C**). For Omicron breakthrough infections following vaccination, geometric mean PRNT_90_ titers of 913.5 (95%CI:341.8-2442) were observed for Omicron, and 627.9 (95%CI:269.5-1463) were observed for Delta, which equals a 1.5-fold loss of neutralization to heterologous Delta (**Fig. 2 D**). Highest titers were seen against the infecting variant Omicron compared to Delta, although the prior vaccine-derived immunity by an early pandemic spike is antigenically closer to Delta than to Omicron.

A heatmap of fold-change reduction of neutralization was performed to summarize findings across all post-vaccine and combined infection/vaccine specimens (**Fig. 2E**). The pronounced escape from vaccination specific for Omicron is visible here, while much less immune escape was observed for the other VOCs as well as for Zeta. Neutralization for Omicron is improved in all specimens with combined infection/vaccination immunity.

We also performed a mapping of our titration results using antigenic cartography (**Fig. 3**). Here we could show that homologous sera cluster around the respective infecting virus, with Alpha and pre-VOC specimens clustering together most closely. Earlier variants of concern before Omicron (Alpha, Beta, Gamma, Delta) belong to one antigenic cluster. Zeta and Omicron are more distantly represented in the map with more than 3 units distance to all other viruses, thus presenting two separate antigenic clusters (**Fig. 3A**). Post vaccine sera cluster around B.1 and Alpha strains, and a larger but equal distance to Beta and Gamma is observed (**Fig. 3B**). Again, Zeta shows a larger antigenic distance, but Omicron is the most distant. Of note, specimens with combined immunity from infection followed by vaccination are located closer to the other non-Alpha VOCs compared to post-vaccination samples without prior infection.

**Figure 3.**
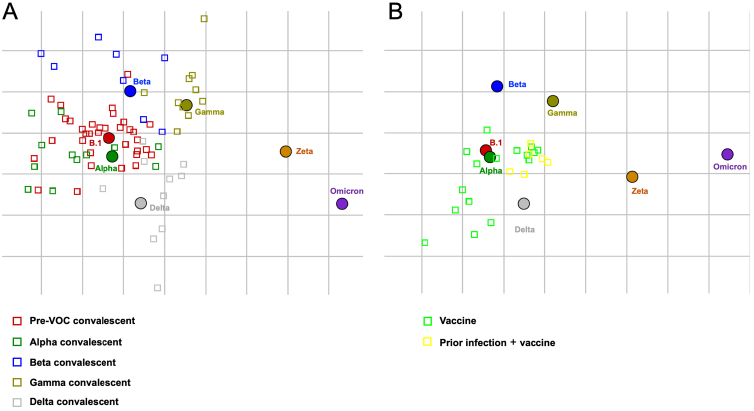
Mapping of antigenic relationship using antigenic cartography for (**A**) convalescent specimens from pre-VOC SARS-CoV-2, Alpha, Beta, Gamma and Delta infections and (**B**) all convalescent and post-vaccination specimens with and without prior infection. For graphical reasons only post vaccination specimens are shown in (**B**)

### Individual profiles of SARS-CoV-2 homologous and heterologous neutralization

In addition to cumulated data (**Fig. 1 and 2**), neutralization profiles were also displayed on an individual basis (**Figure S1-S9**). Here, inter-individual differences both in quantity of antibody response as well as patterns of neutralization loss towards heterologous variants were observed, which does not necessarily reflect pooled results in all individuals. For example, while neutralization against Omicron was the lowest throughout convalescent samples, the differences between Alpha, Beta and Gamma were less pronounced in some individuals. Of note, in all individual comparisons, the most efficient neutralization was always observed against the homologous (infecting) strain through all convalescent sera as well as vaccine sera, when considering that the vaccine Spike most closely resembles the early pandemic variant B.1.

## Discussion

Here we assess neutralizing capacity towards seven SARS-CoV-2 variants by convalescent specimens from individuals recovered from infection with the early-pandemic strain (pre-VOC) or the VOCs Alpha, Beta, Gamma or Delta, from double-vaccinated individuals either with or without prior infection and from double-vaccinated individuals infected with the Delta or Omicron variant.

We could show that highest SARS-CoV-2 neutralizing titers, either elicited through infection of vaccination, were always observed against the homologous strain (infecting strain or antigen used in the vaccine formulation), whereas a reduced neutralizing capacity was found for heterologous strains. In agreement with our results, one study found lower neutralization for the Alpha and Gamma variant by pre-VOC convalescent sera and lower neutralization for pre-VOC and Alpha in Gamma infected patients, but reduced neutralization only for Gamma in vaccine sera [26]. In line with these observations, Alpha-infected individuals showed reduced neutralization for the Beta variant [27]. Reduction in neutralization capacity for the Delta variant was estimated between 4-and 8-fold in vaccinated and 6-fold in convalescent sera [28]. Beyond our study, there is one pre-print that investigated the response to VOCs in a background of variant-specific convalescent samples from individuals infected with pre-VOC, Alpha, Beta or Delta samples, that showed strongest neutralization towards homologous vs heterologous virus for Alpha, Beta and Delta convalescent-variant pairs. [29]

We observed a reduction of neutralization capacity by first-wave SARS-CoV-2 convalescent and post-vaccine samples towards Alpha, Beta, Gamma and Delta to a comparable extent than what was described by others [28, 30-33]. In comparison with neutralizing activity of other convalescent samples, pre-VOC specimens induced immunity showed rather robust neutralization against Alpha, Beta, Gamma and Delta, while convalescent specimens from VOC infections showed lower potential to neutralize heterologous viruses. While for the Alpha variant, only slightly reduced neutralization was described for both convalescent and vaccinee sera, a more pronounced reduction of neutralization was observed for the Beta and Gamma variant [31, 32, 34-37]. Thus, Beta and Gamma comprised the two VOCs with the most pronounced immune escape and at the same time successful enough to cause larger outbreaks in defined geographic region, without reaching global dominance in the time before the emergence of Omicron. A recent study with pre-VOC SARS-CoV-2, Alpha and Beta in multiple animal and cell culture models showed enhanced fitness of Alpha but not Beta which could be a reason why Alpha but not Beta reached wide dominance in early 2021 [22].

Across our panel of convalescent and vaccine sera, the strongest decline of neutralization capacity was observed for Zeta and Omicron. While convalescent specimens showed strong reduction of neutralization for Zeta of up to 25.9-fold, neutralization capacity was restored in vaccinated individuals that showed only 4.3-fold decline compared to pre-VOC SARS-CoV-2. In contrast, for Omicron, a stark loss of neutralizing activity of up to 46-fold or 86-fold was observed for both convalescent and vaccine sera, respectively. Thus, fold-change reduction was even higher in vaccine sera than in convalescent sera, although titer in vaccine sera were higher and therefore percentage of specimens with complete loss was lower (31% of specimens vs. 83%). A similar loss of neutralization capacity against Omicron was observed in other studies [29, 38-40], although no validation across the full range of VOC convalescent sera was available before our data set. Interestingly, strongest loss of neutralization for Zeta was in a similar range to the weakest loss of neutralization for Omicron, e.g. a 25.9-fold reduction of Zeta in pre-VOC convalescent samples, and a 23.0-fold reduction of Omicron in Beta convalescent samples. Such observed differences in fold-change reduction between Zeta and Omicron were less than 2-fold in pre-VOC, Alpha and Beta convalescent sera, showing that even before the emergence of Omicron, variants with strong immune escaping properties almost reaching that of Omicron were already circulating but did not become dominant. For post-Gamma and post-Delta specimens, the differences between fold-change reduction of Zeta and Omicron were much bigger, with Omicron showing a 3-5-fold higher escape of neutralization compared to Zeta. At least for Gamma, robust neutralization for Zeta could be explained by a common origin as both are descendants from B.1.1.28 (Pango lineage) [10]. To the best of our knowledge, no data on neutralization of the Zeta variant by convalescent or vaccinee samples has been described to date, and few other data were obtained on this variant. Contrary, loss of neutralization for Delta was much more pronounced in Gamma than in any other convalescent or vaccine sera with a 17.6-fold reduction. Such strong immune escape of Delta in a variant-specific infection background has not been reported before.

A recent preprint mapped antigenic diversity of SARS-CoV-2, a method that was originally developed to map antigenic relationship for influenza viruses by hemagglutinin inhibition assay [25, 41]. When used to assess influenza viruses, one unit represents a 2-fold change in neutralization titer, and a distance of more than 3 units is required for a separate an antigenic cluster, while below 3, it is considered antigenically similar. They show that ancestral SARS-CoV-2, Alpha, Beta, Gamma and Delta form one antigenic cluster, while Omicron forms a separate antigenic cluster. Differences to our study was that neutralization in this study was only done with a pseudovirus assay and with a lower number of convalescent samples from patients infected with VOCs. Upon analysis for antigenic cartography, our data confirm the findings of Omicron as another antigenic cluster by full virus PRNT, but in addition for the first time we could also show Zeta forms a separate antigenic cluster [25]. Furthermore, we can show that combined immunity after infection/vaccination shows a reduced distance to heterologous variants such as Beta, Gamma and Delta in the map when compared to vaccine samples without additional infection, indicative of broader neutralization.

Of note, we have observed large differences in the reduction of neutralization capacity across specimens with different infection background which has not been described in this detail before. While convalescent specimens of individuals infected with a pre-VOC SARS-CoV-2 and Alpha showed 45.6- and 56.1-fold reduction of neutralization capacity for Omicron, patients previously infected with the Beta, Gamma or Delta variant showed a lower reduction in neutralization capacity of 23.0, 26.9 and 23.8-fold, and a lower percentage of specimens with complete neutralization failure, respectively. These differences could indicate that regional heterogeneity in background immunity could potentially influence emergence and spread of Omicron or other future variants with immune escape properties.

A gradient of immune escape can be described from the variants investigated here, from pre-VOC to Alpha (only slight immune escape, but successfully outcompeted earlier strains), Beta and Gamma (more pronounced immune escape with regionally pronounced circulation but no global dominance) to Zeta (variant with one of the strongest escapes of immunity prior to Omicron, but limited transmission). Our data on Zeta, along with data two other VOIs Mu and Lambda, have shown the strongest escape from neutralization in pandemic period before the emergence of Omicron [42-45]. Although differences between escape from neutralization between Zeta and Omicron are only two-fold in some subgroups, neutralization for Zeta was restored in vaccinee specimens, while this was not the case for Omicron. This could hint towards different mechanisms of immune escape between SARS-CoV-2 variants, and other fitness advantaged for Omicron beyond immune escape.

Of note, a mutation of position 484 in the receptor binding domain is found in Beta, Gamma, Zeta (E484K) and Omicron (E484A). The same mutation arose independently also in a lineage of the Alpha variant where it was also associated with escape from neutralizing antibodies [46]. It has been shown that a mutation at position 484 of the spike tends to have the strongest effect on receptor binding and neutralization [47-50]. While the mutation can explain, at least partly, the strong escape from neutralization in Omicron, it has no or only very low influence on ACE2-binding and is therefore most likely not associated with higher transmissibility, although the mechanism of transmissibility in Omicron is not well understood [51].

We have shown that Omicron exhibits a strong escape from neutralization in both convalescent and vaccine sera, although differences in fold change reduction exist depending on prior infection background. However, it has been shown that a third dose of a mRNA vaccine is able to restore neutralizing capacity [52-55]. Similarly, individuals with mixed immunity, i.e. infection prior to double vaccination, or infection after double vaccination, also leads to higher neutralizing capacity towards Omicron [29, 40]. With an increasing number of vaccine breakthrough infections observed during the Delta wave and reports of a large number of vaccine breakthrough infections with Omicron, an assessment of mixed immunity is of huge interest, especially for protection against new variants [56]. In addition, it has been speculated whether the immune response towards an antigenically drifted SARS-CoV-2 variant will be influenced by pre-existing immunity [57].

Here we investigated four individuals with a Delta breakthrough infection after double vaccination and have found high neutralizing titers for Delta. Neutralization capacity for Omicron was markedly reduced, but titers were still higher than titers against other VOCs in double-vaccinated individuals indicating that even a boost with a mismatched strain (Delta) can lead to a considerable increase in immunity against Omicron. Similarly, vaccine breakthrough infections with Omicron resulted in very high neutralization titers against the Omicron variant indicating that infection with an antigenically drifted variant led to a robust Omicron-specific immune response despite the presence of pre-existing immunity against the original pandemic strain. Interestingly, neutralizing titers against the Delta variant were only minimally reduced compared to Omicron indicating that infection with antigenically different variants can boost immunity against variants that are antigenically similar to the vaccine strain.

Population immunity and evolutionary pressure might differ on a regional scale depending on the immunity induced by earlier circulating variants [58]. High titers of neutralizing antibodies, such as elicited by the vaccine, were at least partially able to neutralize earlier variants before the rise of Omicron. From an epidemiological point of view, this highlights that vaccine-induced immunity probably would have allowed lower virus circulation compared to immunity induced through infections, which may have prevented the evolution of new variants that harbor immune escape properties such as Omicron. Although the origin of Omicron is not well understood, it was speculated that a high background of Beta-variant immunity has favored the development of Omicrons immune escaping properties [59]. In contrast to earlier variants, Omicron is showing a rapid increase in cases with a short doubling time, in addition to its immune-escaping properties [60]. This, together with reduced neutralization is also suggestive of decreased vaccine effectiveness, will further complicate the management of the pandemic.

Limitations of our study are the relatively low number of specimens that were available for convalescent specimens of patients previously infected with variants. Furthermore, we cannot exclude, that individuals infected with a VOC have not already been infected in 2020 with a first-wave virus and thus antibodies are derived from multiple infections with more than one variant. In addition, the Beta variant, which could not be readily isolated on VeroE6 cell lines had to be adapted to VeroE6 in order to be usable for our PRNT, thus the isolate accumulated mutations, which could affect the neutralization results. No convalescent samples were available from individuals infected with Zeta, and no convalescent samples were yet available from individuals infected with Omicron without prior vaccination. Furthermore, testing of single time points after infection/vaccination can only provide a snapshot and not inform on the duration of antibody responses over time. All our blood specimens were collected at rather early time points after infection or vaccination, thus with time, a broader neutralizing capacity towards the heterologous variant might be observed due to somatic hypermutation and affinity maturation, although in the case of Omicron most likely will not restore neutralization loss [61].

Overall, we could show that responses to variants before Omicron were associated with reduced, but not complete loss in neutralization both by infection-derived as well as vaccine derived immunity, although variants such as Zeta have already shown immune escape properties, but did not become dominant at the time of their emergence. Omicron, in contrast to earlier variants displays a strong immune escape with a surge in case numbers in many areas of the world. Furthermore, the continuous emergence of SARS-CoV-2 variants since late 2020 shows that the virus is still underlies evolutionary pressure and currently available vaccines might not be sufficient to mitigate the pandemic in the near-term future. Furthermore, whether immunity of a population was obtained by a vaccine or by previous infection with a certain SARS-CoV-2 variants could influence the type of evolutionary pressure for circulating SARS-CoV-2 and lead to the emergence of new variants that differ for countries with major vaccine vs. major infection-derived immunity.

## Data Availability

All data produced in the present study are available upon reasonable request to the authors

## Acknowledgments

We thank Pascale Sattonnet-Roche for excellent technical help. We thank the staff of the laboratory of virology at the HUG for support. We thank all clinicians and technical staff responsible of the different clinical cohorts for their help. We are grateful for the patients who were willing to donate their samples and agree to participate in our research.

We thank Samuel Cordey and Florian Laubscher for help with sequence analysis. We thank Mirco Schmolke and Beryl Mazel-Sanchez for A549-hACE2 cells.

We thank Volker Thiel, Jenna Kelly and Silvio Steiner, Vetsuisse Bern, for help with Omicron sequencing.

## Supplementary data

**Table S1.** Patient sample information from which virus isolates used in this study were obtained.

|  | **B.1** | **Alpha** | **Beta** | **Gamma** | **Delta** | **Zeta** | **Omicron** |
| --- | --- | --- | --- | --- | --- | --- | --- |
| **Name** | hCoV-19/Switzerland/GE-SNRCI-29943121/2020 | hCoV-19/Switzerland/2012212272/2020 | hCoV-19/Switzerland/GE-33128281/2021 | hCoV-19/Switzerland/GE-33115015/2021 | hCoV-19/Switzerland/GE-33896105/2021 | hCoV-19/Switzerland/GE-32966260/2021 | hCoV-19/Switzerland/VD-HUG-36221084/2021 |
| **GISAID accession ID** | EPI_ISL_414019 | EPI_ISL_2131446 | EPI_ISL_981782 | EPI_ISL_981707 | EPI_ISL_1811202 | EPI_ISL_897700 | EPI_ISL_7605546 |
| **Clade** | G | GRY | GH | GR | GK | G | GRA |
| **Pango lineage** | B.1 | B.1.1.7 | B.1.351 | P.1 | AY.122 | P.2 | BA.1 |

**Figure S1.**
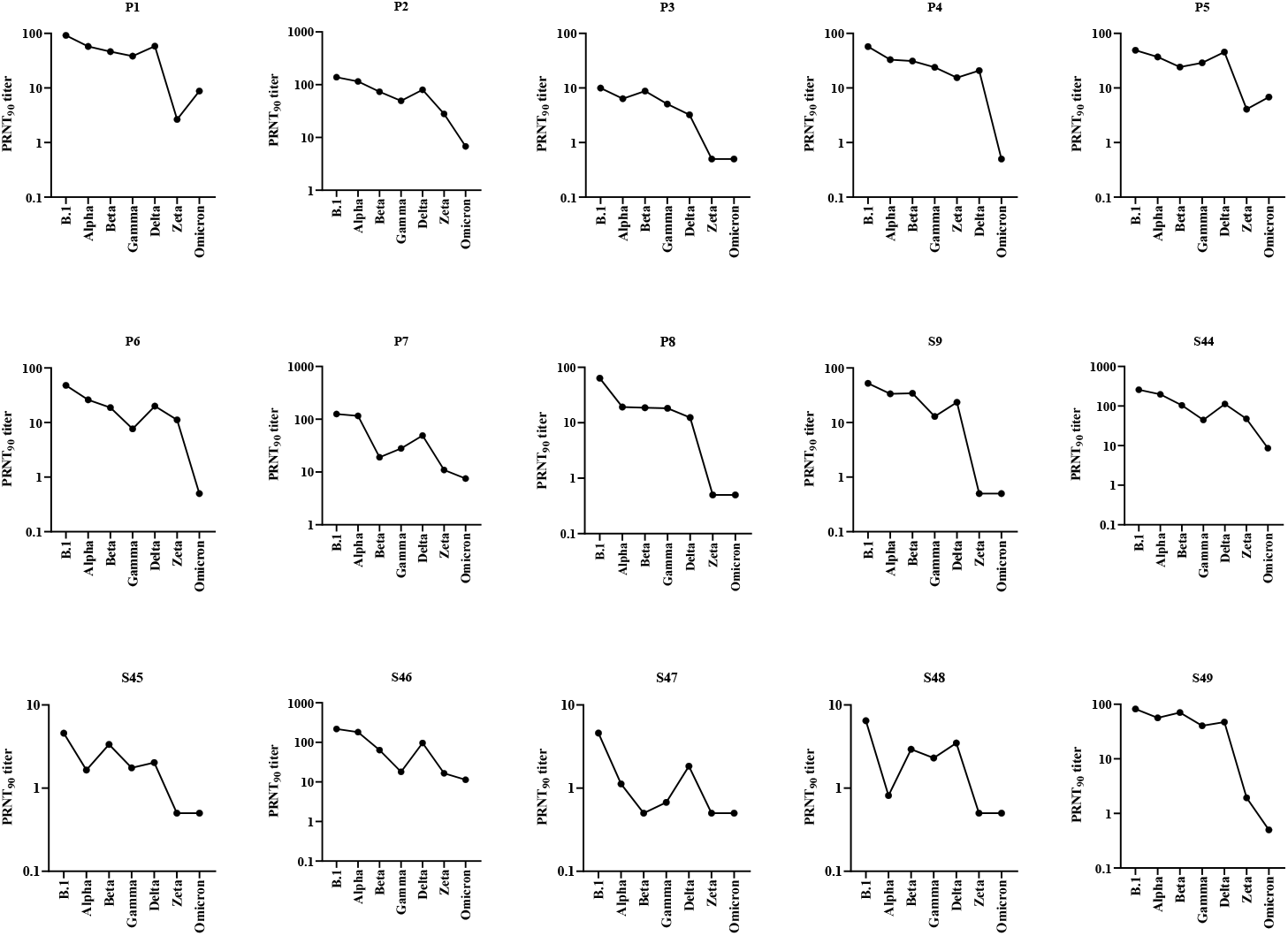
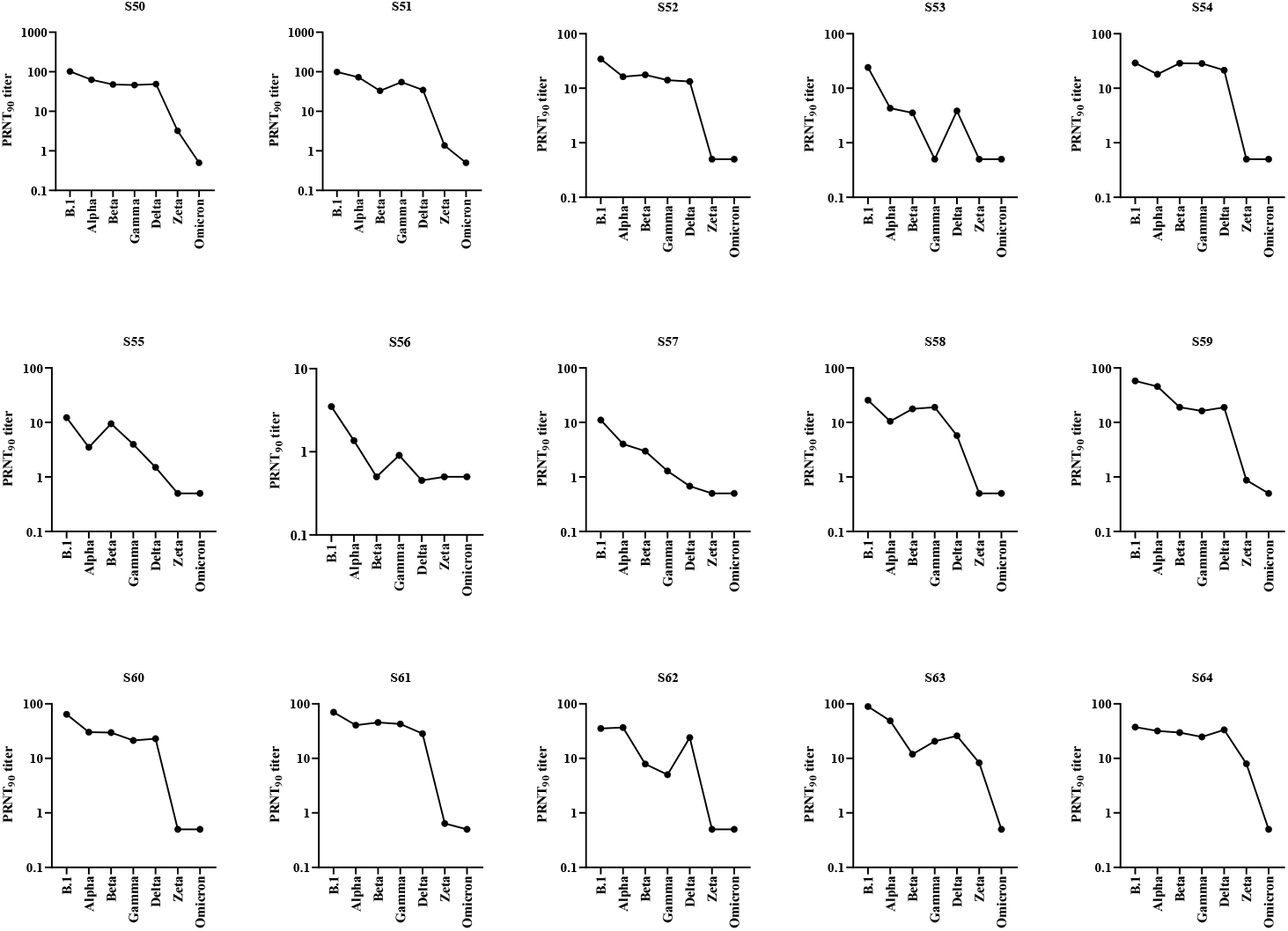
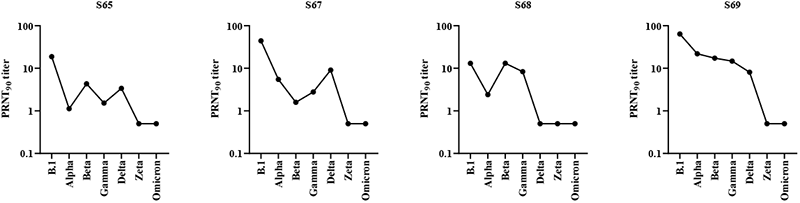
Neutralizing antibody titers of each pre-VOC convalescent individual (P=plasma sample, S= serum sample).

**Figure S2.**
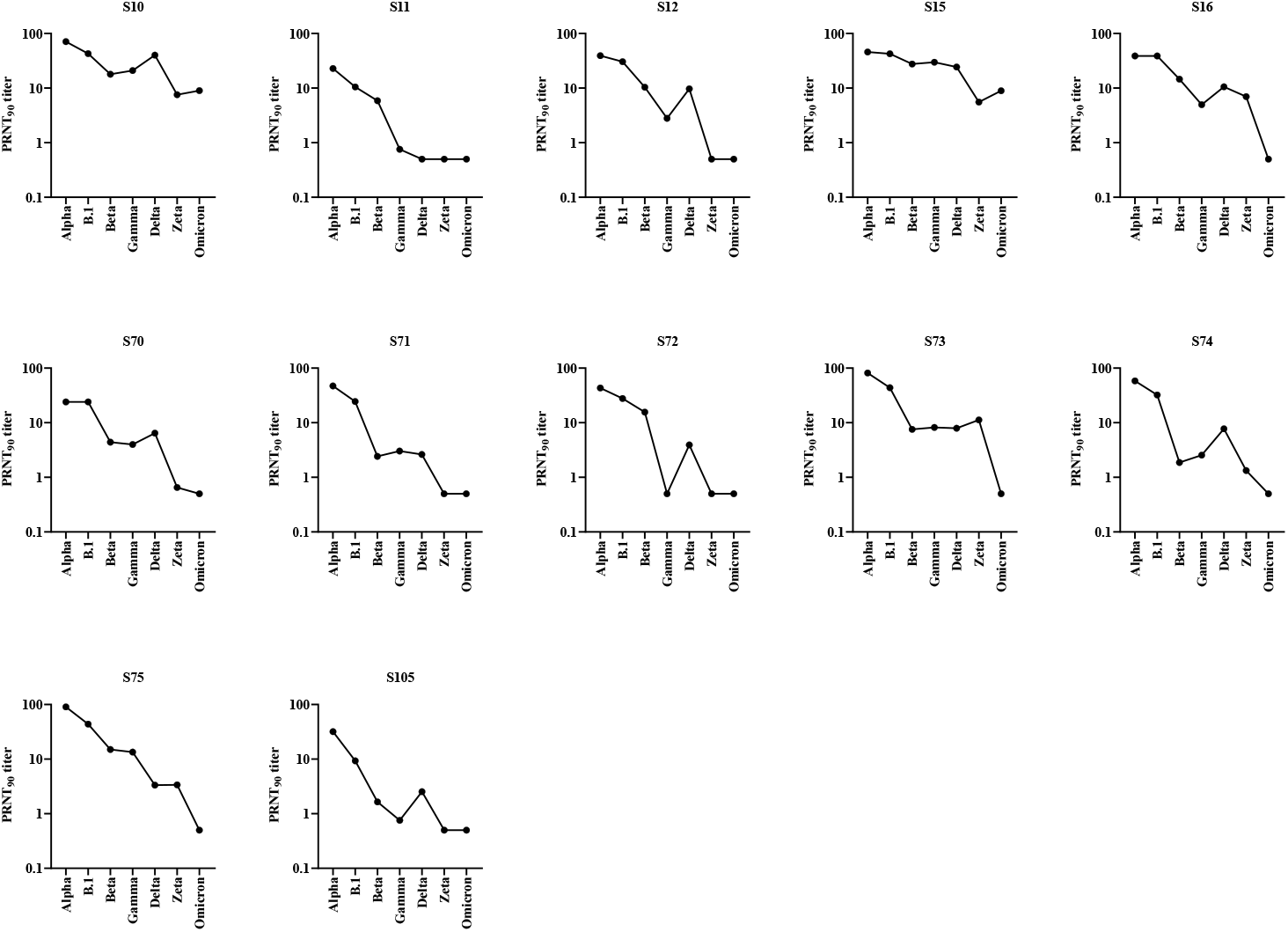
Neutralizing antibody titers of each Alpha convalescent individual (S= serum sample).

**Figure S3.**
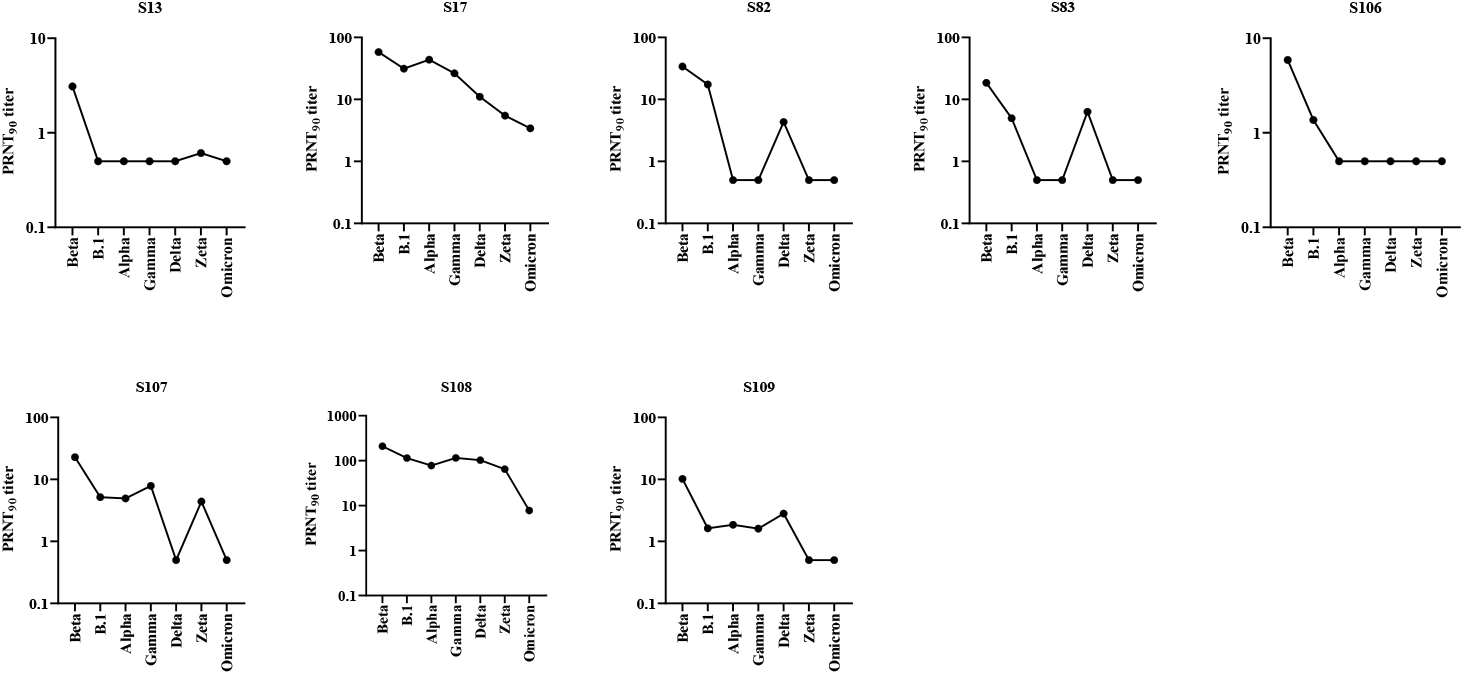
Neutralizing antibody titers of each Beta convalescent individual (S= serum sample).

**Figure S4.**
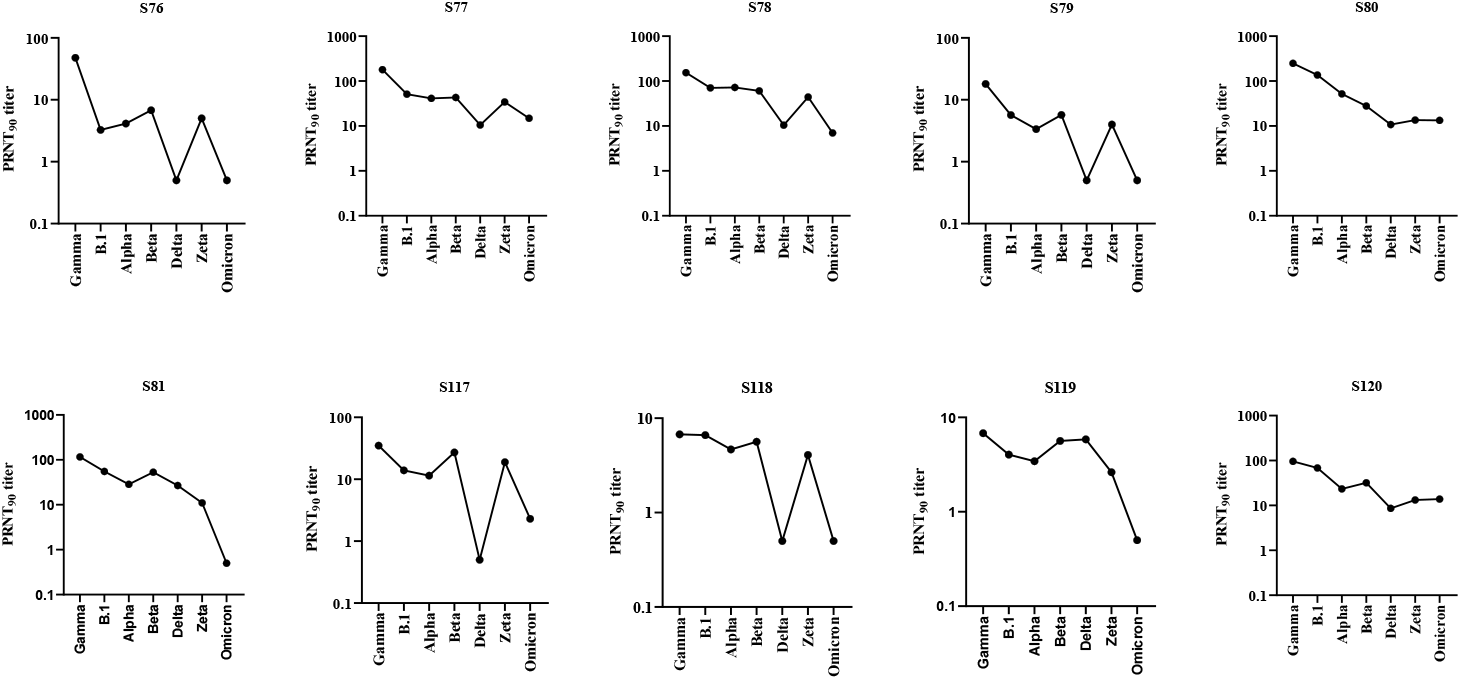
Neutralizing antibody titers of each Gamma convalescent individual (S= serum sample).

**Figure S5.**
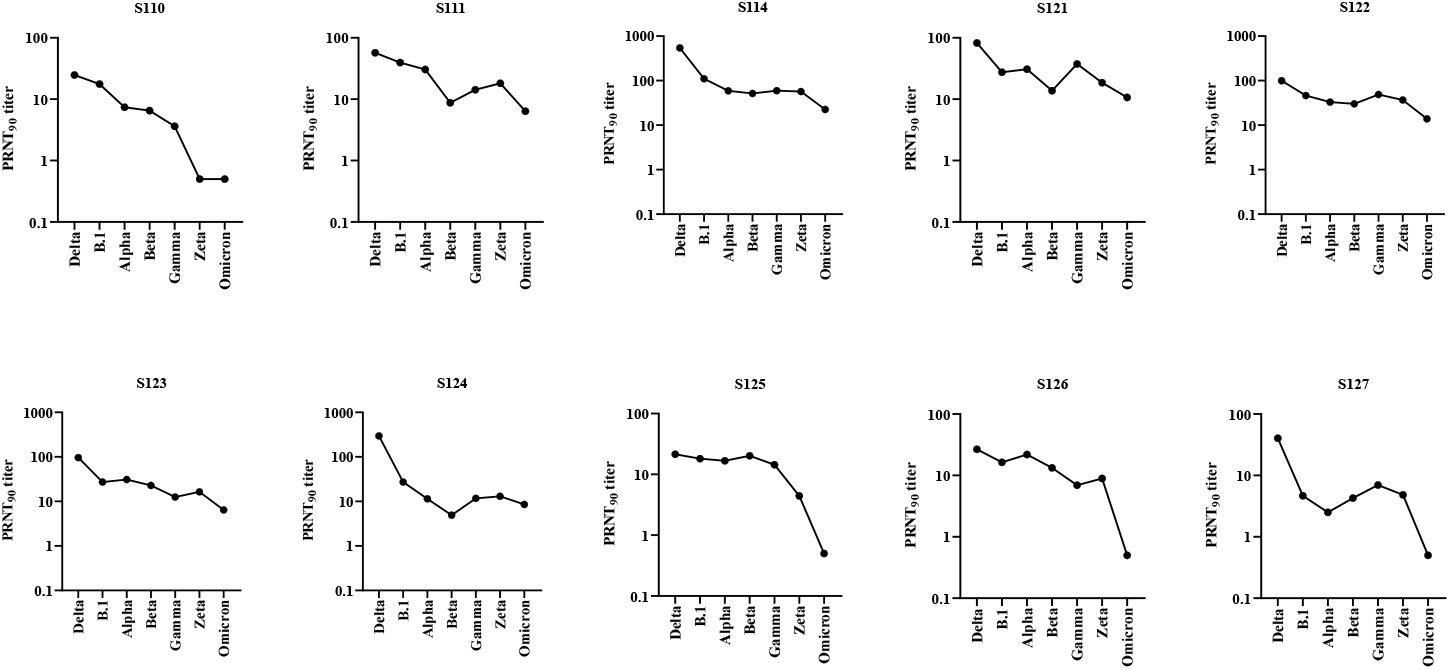
Neutralizing antibody titers of each Delta convalescent individual (S= serum sample).

**Figure S6.**
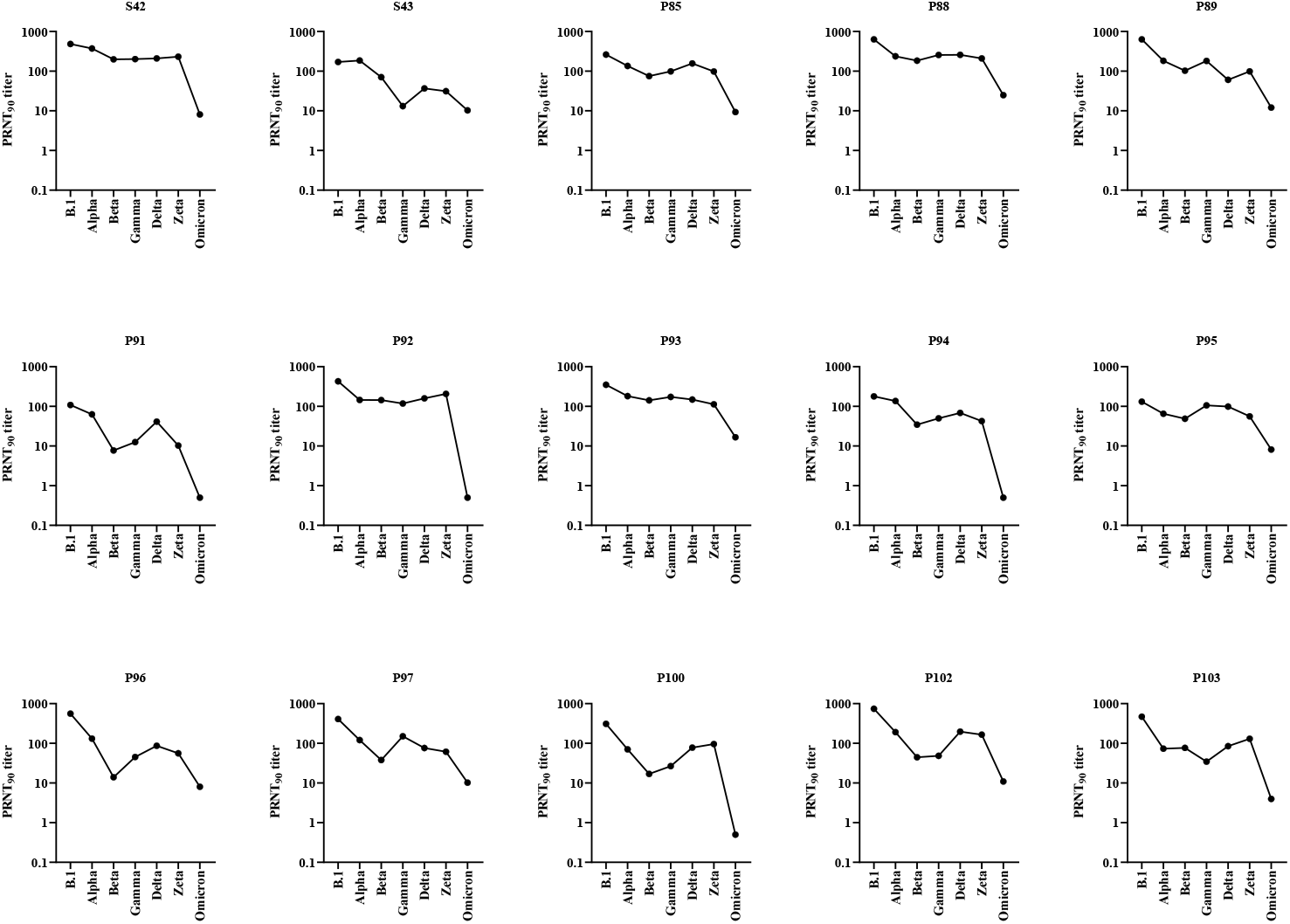
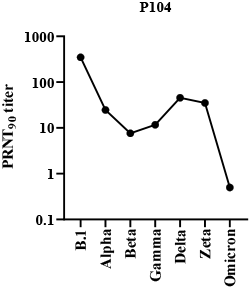
Neutralizing antibody titers of each post-vaccination individual with double vaccination) (P=plasma sample, S= serum sample).

**Figure S7.**
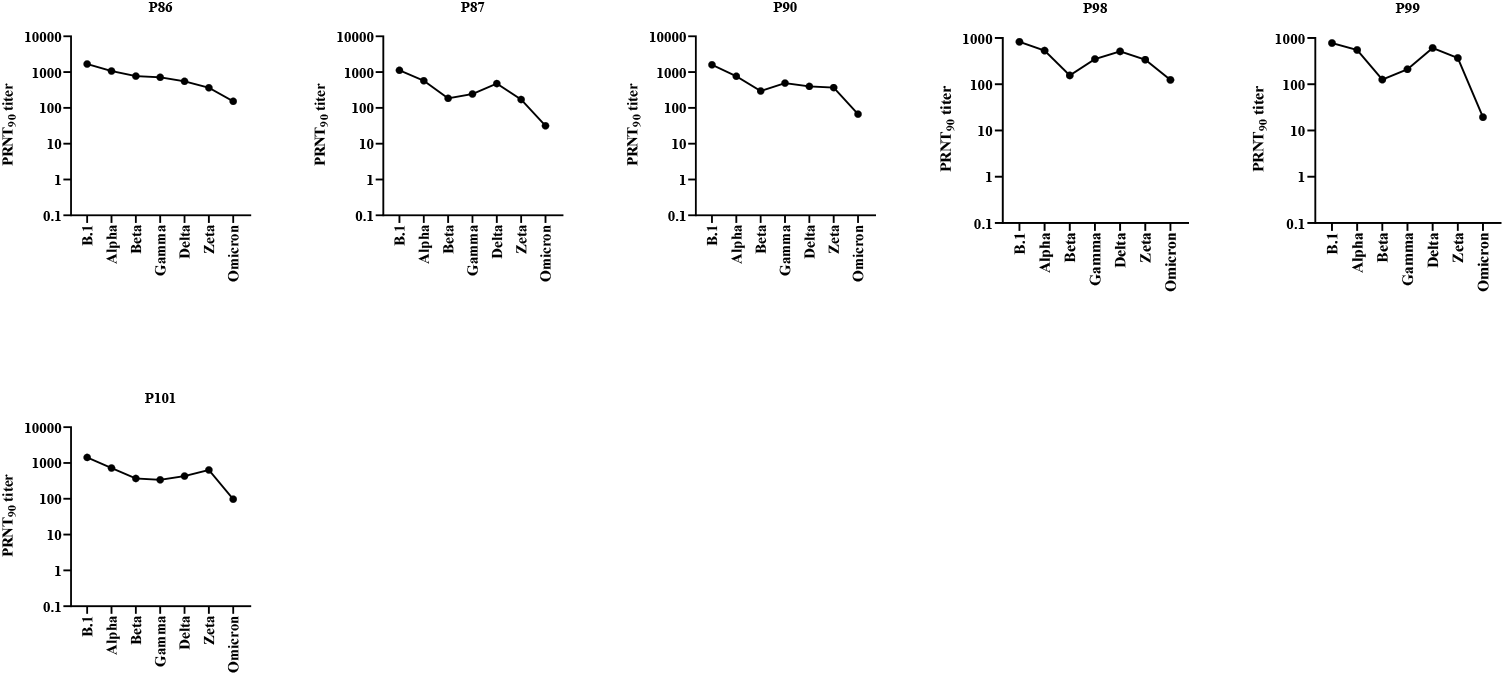
Neutralizing antibody titers of each post-vaccination individual with prior infection followed by double vaccination (P=plasma sample).

**Figure S8.**
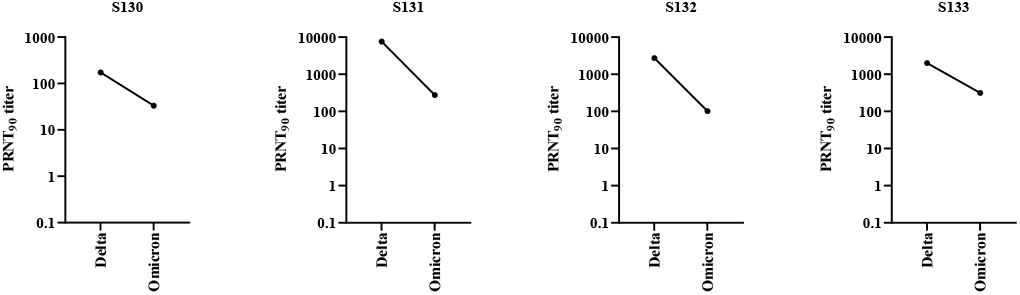
Neutralizing antibody titers of each post-vaccination individual with Delta breakthrough infection (S= serum sample).

**Figure S9.**
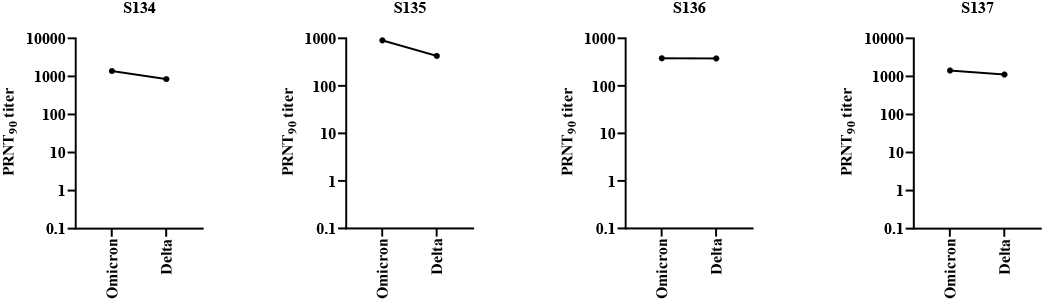
Neutralizing antibody titers of each post-vaccination individual with Omicron breakthrough infection (S= serum sample).

## References

1. Zhu, N., et al., A Novel Coronavirus from Patients with Pneumonia in China, 2019. N Engl J Med, 2020. 382(8): p. 727–733.

2. Zhou, P., et al., A pneumonia outbreak associated with a new coronavirus of probable bat origin. Nature, 2020. 579(7798): p. 270–273.

3. Zhou, B., et al., SARS-CoV-2 spike D614G change enhances replication and transmission. Nature, 2021. 592(7852): p. 122–127.

4. World Health Organization (WHO) 08.12.2021]; Available from: https://www.who.int/en/activities/tracking-SARS-CoV-2-variants/.

5. Viana, R., et al., Rapid epidemic expansion of the SARS-CoV-2 Omicron variant in southern Africa. medRxiv, 2021: p. 2021.12.19.21268028.

6. Tegally, H., et al., Detection of a SARS-CoV-2 variant of concern in South Africa. Nature, 2021. 592(7854): p. 438–443.

7. Davies, N.G., et al., Estimated transmissibility and impact of SARS-CoV-2 lineage B.1.1.7 in England. Science, 2021. 372(6538).

8. Campbell, F., et al., Increased transmissibility and global spread of SARS-CoV-2 variants of concern as at June 2021. Euro Surveill, 2021. 26(24).

9. Faria, N.R., et al., Genomics and epidemiology of the P.1 SARS-CoV-2 lineage in Manaus, Brazil. Science, 2021. 372(6544): p. 815–821.

10. Voloch, C.M., et al., Genomic characterization of a novel SARS-CoV-2 lineage from Rio de Janeiro, Brazil. J Virol, 2021.

11. NIH. 08.12.2021]; Available from: https://www.covid19treatmentguidelines.nih.gov.

12. Walsh, E.E., et al., Safety and Immunogenicity of Two RNA-Based Covid-19 Vaccine Candidates. N Engl J Med, 2020. 383(25): p. 2439–2450.

13. Jackson, L.A., et al., An mRNA Vaccine against SARS-CoV-2 -Preliminary Report. N Engl J Med, 2020. 383(20): p. 1920–1931.

14. Wang, R., et al., Vaccine-escape and fast-growing mutations in the United Kingdom, the United States, Singapore, Spain, India, and other COVID-19-devastated countries. Genomics, 2021. 113(4): p. 2158–2170.

15. Hoffmann, M., et al., SARS-CoV-2 variants B.1.351 and P.1 escape from neutralizing antibodies. Cell, 2021. 184(9): p. 2384–2393 e12.

16. Addetia, A., et al., Neutralizing Antibodies Correlate with Protection from SARS-CoV-2 in Humans during a Fishery Vessel Outbreak with a High Attack Rate. J Clin Microbiol, 2020. 58(11).

17. Khoury, D.S., et al., Neutralizing antibody levels are highly predictive of immune protection from symptomatic SARS-CoV-2 infection. Nat Med, 2021. 27(7): p. 1205–1211.

18. Earle, K.A., et al., Evidence for antibody as a protective correlate for COVID-19 vaccines. Vaccine, 2021. 39(32): p. 4423–4428.

19. Huang, A.T., et al., A systematic review of antibody mediated immunity to coronaviruses: antibody kinetics, correlates of protection, and association of antibody responses with severity of disease. medRxiv, 2020.

20. Chmielewska, A.M., et al., Immune response against SARS-CoV-2 variants: the role of neutralization assays. NPJ Vaccines, 2021. 6(1): p. 142.

21. Federal Office for Public Health Switzerland 08.12.2021]; Available from: https://www.bag.admin.ch/bag/en/home/das-bag/aktuell/medienmitteilungen.msg-id-83732.html.

22. Ulrich, L., et al., Enhanced fitness of SARS-CoV-2 variant of concern Alpha but not Beta. Nature, 2021.

23. Ilmjarv, S., et al., Concurrent mutations in RNA-dependent RNA polymerase and spike protein emerged as the epidemiologically most successful SARS-CoV-2 variant. Sci Rep, 2021. 11(1): p. 13705.

24. Meyer, B., et al., Validation of a commercially available SARS-CoV-2 serological immunoassay. Clin Microbiol Infect, 2020. 26(10): p. 1386–1394.

25. van der Straten, K., et al., Mapping the antigenic diversification of SARS-CoV-2. medRxiv, 2022: p. 2022.01.03.21268582.

26. Gidari, A., et al., Cross-neutralization of SARS-CoV-2 B.1.1.7 and P.1 variants in vaccinated, convalescent and P.1 infected. J Infect, 2021. 83(4): p. 467–472.

27. Faulkner, N., et al., Reduced antibody cross-reactivity following infection with B.1.1.7 than with parental SARS-CoV-2 strains. Elife, 2021. 10.

28. Davis, C., et al., Reduced neutralisation of the Delta (B.1.617.2) SARS-CoV-2 variant of concern following vaccination. PLoS Pathog, 2021. 17(12): p. e1010022.

29. Rössler, A., et al., SARS-CoV-2 B.1.1.529 variant (Omicron) evades neutralization by sera from vaccinated and convalescent individuals. medRxiv, 2021: p. 2021.12.08.21267491.

30. Hojjat Jodaylami, M., et al., Cross-reactivity of antibodies from non-hospitalized COVID-19 positive individuals against the native, B.1.351, B.1.617.2, and P.1 SARS-CoV-2 spike proteins. Sci Rep, 2021. 11(1): p. 21601.

31. Muik, A., et al., Neutralization of SARS-CoV-2 lineage B.1.1.7 pseudovirus by BNT162b2 vaccine-elicited human sera. Science, 2021. 371(6534): p. 1152–1153.

32. Shen, X., et al., Neutralization of SARS-CoV-2 Variants B.1.429 and B.1.351. N Engl J Med, 2021. 384(24): p. 2352–2354.

33. Mlcochova, P., et al., SARS-CoV-2 B.1.617.2 Delta variant replication and immune evasion. Nature, 2021. 599(7883): p. 114–119.

34. Edara, V.V., et al., Infection- and vaccine-induced antibody binding and neutralization of the B.1.351 SARS-CoV-2 variant. Cell Host Microbe, 2021. 29(4): p. 516–521 e3.

35. Skelly, D.T., et al., Two doses of SARS-CoV-2 vaccination induce robust immune responses to emerging SARS-CoV-2 variants of concern. Nat Commun, 2021. 12(1): p. 5061.

36. Wang, P., et al., Increased resistance of SARS-CoV-2 variant P.1 to antibody neutralization. Cell Host Microbe, 2021. 29(5): p. 747–751 e4.

37. Bates, T.A., et al., Age-Dependent Neutralization of SARS-CoV-2 and P.1 Variant by Vaccine Immune Serum Samples. JAMA, 2021.

38. Dejnirattisai, W., et al., Reduced neutralisation of SARS-COV-2 Omicron-B.1.1.529 variant by post-immunisation serum. medRxiv, 2021: p. 2021.12.10.21267534.

39. Cele, S., et al., SARS-CoV-2 Omicron has extensive but incomplete escape of Pfizer BNT162b2 elicited neutralization and requires ACE2 for infection. medRxiv, 2021: p. 2021.12.08.21267417.

40. Wilhelm, A., et al., Reduced Neutralization of SARS-CoV-2 Omicron Variant by Vaccine Sera and monoclonal antibodies. medRxiv, 2021: p. 2021.12.07.21267432.

41. Smith, D.J., et al., Mapping the antigenic and genetic evolution of influenza virus. Science, 2004. 305(5682): p. 371–6.

42. Acevedo, M.L., et al., Infectivity and immune escape of the new SARS-CoV-2 variant of interest Lambda. medRxiv, 2021: p. 2021.06.28.21259673.

43. Xie, X., et al., Emerging SARS-CoV-2 B.1.621/Mu variant is prominently resistant to inactivated vaccine-elicited antibodies. Zool Res, 2021. 42(6): p. 789–791.

44. Uriu, K., et al., Neutralization of the SARS-CoV-2 Mu Variant by Convalescent and Vaccine Serum. N Engl J Med, 2021. 385(25): p. 2397–2399.

45. Liu, H., et al., The Lambda variant of SARS-CoV-2 has a better chance than the Delta variant to escape vaccines. bioRxiv, 2021.

46. Collier, D.A., et al., Sensitivity of SARS-CoV-2 B.1.1.7 to mRNA vaccine-elicited antibodies. Nature, 2021. 593(7857): p. 136–141.

47. Greaney, A.J., et al., Comprehensive mapping of mutations in the SARS-CoV-2 receptor-binding domain that affect recognition by polyclonal human plasma antibodies. Cell Host Microbe, 2021. 29(3): p. 463–476 e6.

48. Andreano, E., et al., SARS-CoV-2 escape in vitro from a highly neutralizing COVID-19 convalescent plasma. bioRxiv, 2020.

49. Liu, Z., et al., Landscape analysis of escape variants identifies SARS-CoV-2 spike mutations that attenuate monoclonal and serum antibody neutralization. bioRxiv, 2021.

50. Weisblum, Y., et al., Escape from neutralizing antibodies by SARS-CoV-2 spike protein variants. Elife, 2020. 9.

51. Starr, T.N., et al., Deep Mutational Scanning of SARS-CoV-2 Receptor Binding Domain Reveals Constraints on Folding and ACE2 Binding. Cell, 2020. 182(5): p. 1295–1310 e20.

52. Garcia-Beltran, W.F., et al., mRNA-based COVID-19 vaccine boosters induce neutralizing immunity against SARS-CoV-2 Omicron variant. medRxiv, 2021.

53. Doria-Rose, N.A., et al., Booster of mRNA-1273 Vaccine Reduces SARS-CoV-2 Omicron Escape from Neutralizing Antibodies. medRxiv, 2021.

54. Nemet, I., et al., Third BNT162b2 vaccination neutralization of SARS-CoV-2 Omicron infection. medRxiv, 2021: p. 2021.12.13.21267670.

55. Yu, X., et al., Enhanced neutralization against SARS-CoV-2 by vaccine booster exhibits reduction of Omicron variant. medRxiv, 2021: p. 2021.12.17.21267961.

56. Eggink, D., et al., Increased risk of infection with SARS-CoV-2 Omicron compared to Delta in vaccinated and previously infected individuals, the Netherlands, 22 November to 19 December 2021. medRxiv, 2021: p. 2021.12.20.21268121.

57. Brown, E.L. and H.T. Essigmann, Original Antigenic Sin: the Downside of Immunological Memory and Implications for COVID-19. mSphere, 2021. 6(2).

58. Karuna, S., et al., Neutralizing antibody responses over time in demographically and clinically diverse individuals recovered from SARS-CoV-2 infection in the United States and Peru: A cohort study. PLoS Med, 2021. 18(12): p. e1003868.

59. Zahradnik, J., et al., Receptor binding and escape from Beta antibody responses drive Omicron-B.1.1.529 evolution. bioRxiv, 2021: p. 2021.12.03.471045.

60. Yang, W. and J. Shaman, SARS-CoV-2 transmission dynamics in South Africa and epidemiological characteristics of the Omicron variant. medRxiv, 2021: p. 2021.12.19.21268073.

61. Gaebler, C., et al., Evolution of antibody immunity to SARS-CoV-2. Nature, 2021. 591(7851): p. 639–644.

